# Analysis of the response of prostate cancer to ultra-hypofractionated high-dose-rate brachytherapy: the role of hypoxia and reoxygenation

**DOI:** 10.64898/2026.05.07.26352634

**Authors:** Eva G. Kölmel, Pedro Otero-Casal, Juan Pardo-Montero

## Abstract

Clinical studies of prostate cancer treated with radically hypofractionated highdose-rate brachytherapy (HDR-BT) have reported a significant loss of tumor control that contradicts the standard linear-quadratic (LQ) and low *α/β* ratio paradigm for prostate cancer. In a previous study by our group, we showed that the linear-quadratic-linear (LQL) model could describe this response, but the underlying biological drivers remained unclear. In this follow-up study, we further investigate whether the interplay between hypoxia and reoxygenation kinetics can explain the poor response to extreme hypofractionation. We analyzed a large dataset of 3,239 patients (44 schedules) using a three-compartment reoxygenation model (the MSK model) that simulates the dynamics of oxic, intermediate, and hypoxic cell populations. Results show that the MSK model achieves an excellent fit to the clinical data (*p* > 0.99) while maintaining a biologically plausible low *α/β* ratio (*≤* 8 Gy). The reoxygenation model provided a performance comparable to the LQL model for low-risk prostate cancer, being slightly inferior to the LQL model to describe the response of intermediate-risk. This suggests that the observed reduction in tumor control is not necessarily a failure of the LQ formalism, but rather a consequence of oxygen dynamics associated with ultra-fractionated schedules, and provides a mechanistic basis for designing clinical trials exploring the response of prostate cancer to ultra-hypofractionation and the role of reoxygenation.

## 1 Introduction

The response of prostate cancer to radiotherapy is probably the most studied case in the radiobiological modeling literature [1, 2, 3]. The clear consensus is that the *α/β* ratio of prostate cancer is very low (typically in the 1–4 Gy range), and this tumor is very sensitive to fractionation. In recent years, extreme hypofractionation has become widely used to treat prostate cancer, by using either stereotactic body radiotherapy (SBRT) or high-dose-rate brachytherapy (HDR-BT). While SBRT treatments reach 10 Gy per fraction, HDR-BT treatments can reach >20 Gy in a single fraction. Interestingly, those ultra-hypofractionation studies have reported a significant reduction in tumor control (<70%), which seems to be in contradiction with the classical paradigm of a low *α/β* ratio for prostate cancer.

Guirado *et al*. were the first group to analyze the response of prostate cancer to hypofractionated HDR-BT, proposing a large *α/β* ratio (∼23 Gy) to explain the poor control achieved with single-fraction HDR-BT treatments [4]. This value is in conflict with many radiobiological studies on *classical* radiotherapy and SBRT that support a low *α/β* for prostate cancer. Therefore, they also argued that the linear-quadratic (LQ) model may not be adequate for very large doses per fraction. The latter proposal was further explored in a recent publication from our group [5]. We curated a large dataset of response to HDR-BT including multiple fractionation schemes, and analyzed it with different models, showing that the LQL model of dose-response [6], which softens the damage predicted by the LQ model at large doses per fraction, was capable of describing the clinical response to single-fraction HDR-BT treatments while keeping a low *α/β* ratio.

While the LQ model has been long questioned [7], and may be behind the poor response observed for single-fraction HDR-BT, other biophysical factors could also play a role. In particular, reoxygenation (or the lack thereof for hypofractionated schemes) has been investigated in the context of prostate cancer, starting with the seminal work by Nahum *et al*. [8] (aiming at conventional radiotherapy), and could play a role in the poor response to single-fraction HDR-BT. If reoxygenation and radiosensitization play an important role in the response to fractionated radiotherapy of otherwise hypoxic tumors, the lack thereof associated with extreme hypofractionation may explain the observed poor control.

In Ref. [5] we also performed a preliminary analysis of the effect of reoxygenation by using a simple LQ+reoxygenation model [9], but found no improvement over the simple LQ model when including reoxygenation. In this work, we have extended the analysis of the role of hypoxia and reoxygenation by using a more complex and realistic model originally introduced by Jeong *et al*. [10], which has been well validated to analyze dose-response [11, 12]. This study constitutes a stepping stone towards understanding the mechanisms underlying the response to ultra-fractionated radiotherapy. A deep understanding of such mechanisms seems of paramount importance to design effective ultra-fractionated treatments.

## 2 Materials and Methods

### 2.1 Clinical dataset

Building upon our previous work [5], this study uses the same clinical dataset, which allows for a systematic intercomparison with models analyzed in that work (LQ, LQL and Stavrev *et al*. reoxygenation). The dataset contains dose-response data (local control at five years) for 21 schedules (1633 patients) for low-risk (LR) and 23 schedules (1606 patients) for intermediate-risk (IR) prostate cancer, with doses per fraction ranging from 6 Gy to 21 Gy.

### 2.2 Radiobiological modelling

#### 2.2.1 MSK model

In this work we have adapted the response model of Jeong *et al*. [10] to analyze the clinical response of prostate cancer to HDR-BT. We will refer to this model as the MSK model, in reference to the institution of the authors. This model is a compartmental model which splits tumor cells into three different compartments according to their oxygenation levels: (P) well-oxygenated cells (which are proliferative), (I) intermediate-oxygenated cells, and (H) hypoxic cells. Radiation damage is modeled using the LQ model and affects cells differently according to their oxygenation status (well oxygenated cells being the most sensitive to radiation due to the oxygen enhancement ratio, OER). Damaged cells die when trying to undergo mitosis in the P compartment, which creates vacancies in the P compartment that trigger a shift from poorly oxygenated to well-oxygenated compartments, thereby reoxygenating the tumor. This model has been validated with clinical data [11, 12]. In particular, in Ref. [11] the authors used the model to describe the dose-response of a large dataset of NSCLC.

For further details, we refer the reader to the original publication, but for completeness we present the main equations of the model here. 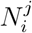 refers to cells in state *i* belonging to the *j* compartment, with *i*=(v, d) referring to viable (v) and doomed due to radiation damage (d), and *j*=(P, I, H) referring to the proliferative (oxic), intermediate, and hypoxic compartments, respectively. In differential form, the model can be written as:

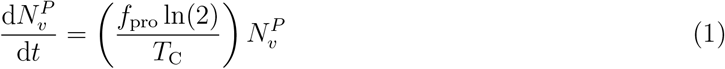

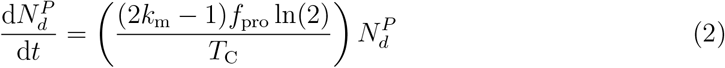

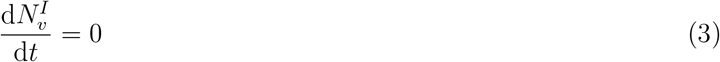

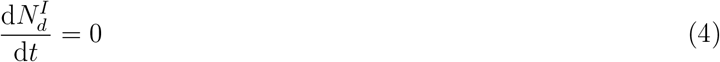

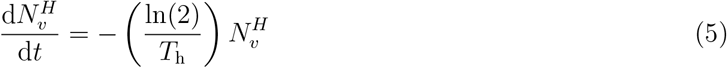

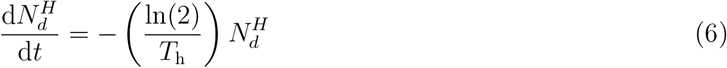

Notice that cells in the I compartment have no dynamics, as they cannot proliferate and do not die during mitosis (they only do so if/when they move to the proliferative compartment). Viable cells in the P compartment proliferate, and doomed cells in P die during mitosis (notice that (2*k*_m_ − 1) < 0). Cells in the H compartment die due to severe hypoxia. Each radiation dose acts as an impulse to the differential equations, moving cells from viable to doomed in each compartment for each radiation dose, *d*, delivered at time *tD*, as:

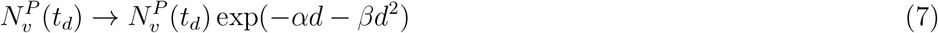

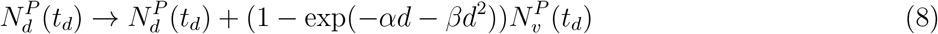

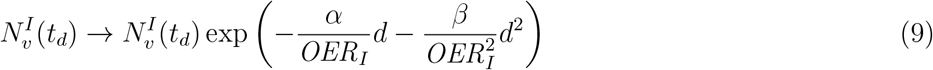

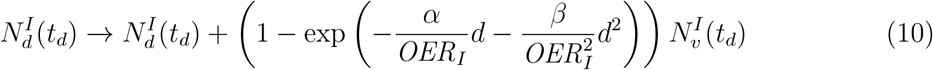

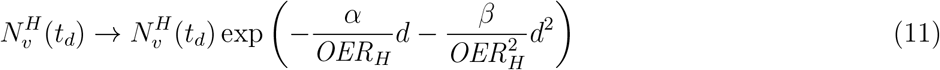

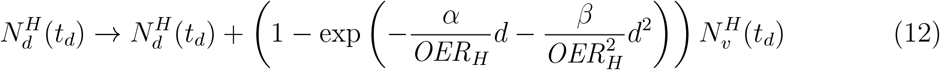

The capacities of compartments P and I are finite and given by:

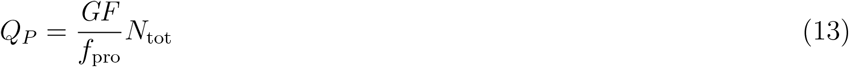

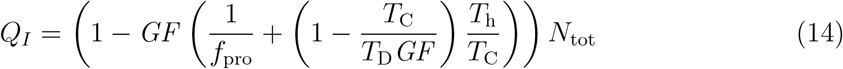

At the beginning of the simulation the tumor contains *N*_tot_ cell, all of which are viable. Cells are assigned to compartment P until it is filled, then to compartment I until filled, and then to compartment H (if necessary). The death of doomed cells in the P compartment creates vacancies which are instantly filled by cells (if any) shifting from the I compartment (both viable and doomed according to their ratio in the I compartment) and the H compartment (if necessary). The shift from I to P creates vacancies in the I compartment that are instantly filled by cells (if any) moving from the H compartment. This compartmental reorganization reoxygenates the tumor.

The model also includes dead cells, which are eventually removed through lysis. However, dead cells do not contribute to the occupancy of the compartments (because they no longer consume oxygen), therefore they do not affect the dynamics of viable and doomed cells or the calculation of tumor control probabilities (the endpoint modeled in this work), and were excluded from this analysis (they would however affect the dynamics of tumor volumes and should be included if modelling that aspect).

A list of the parameters of the model is presented in Table 1.

**Table 1:** Radiobiological parameters of the MSK compartmental model. For parameters that have been set to a fixed value we provide such values (see section 2.3 for more details), while parameters that have been fitted to the experimental data are labelled as *Variable*.

| Parameter | Description | Value (Unit) |
| --- | --- | --- |
| $f_{\text{pro}}$ | Proliferating fraction in the $P$ compartment | Variable |
| $T_C$ | Cell cycle time | 48 h |
| $T_D$ | Tumor doubling time | 34.83 months (LR), 30.43 months (IR) |
| $GF$ | Growth fraction of the tumor | 0.083 (LR), 0.116 (IR) |
| $k_m$ | Mitotic survival probability of doomed cells | Variable |
| $T_h$ | Half-life of cells in the hypoxic compartment $H$ | 48 h |
| $\alpha$ | Linear radiosensitivity parameter for oxic cells | $0.15 \text{ Gy}^{-1}$ |
| $\alpha/\beta$ | $\alpha/\beta$ ratio for oxic cells | Variable |
| $\text{OER}_I$ | Oxygen Enhancement Ratio for compartment $I$ | Variable |
| $\text{OER}_H$ | Oxygen Enhancement Ratio for compartment $H$ | 1.37 |
| $N_{\text{tot}}$ | Total initial number of cells | $10^{10}$ |

### 2.2.2 Tumor control probability and EQD2

The tumor control probability (TCP) was modeled using a logistic function of *equivalent dose in 2 Gy fractions, EQD2*, with parameters *D*_50_ and *γ*_50_ controlling the dose yielding 50% control and the slope of the dose-response curve, respectively:

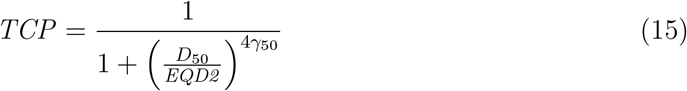

The *equivalent dose in 2 Gy fractions, EQD2*, was numerically obtained from the model, following [11]. To this end, a reference treatment delivering 150 fractions daily of 2 Gy is simulated, and the number of viable cells at the end of each fraction is saved. These values are then interpolated using a spline function. Finally, in order to calculate the *EQD2* of a given schedule, the specific treatment protocol is simulated, and the number of viable cells at the end of the treatment is saved and mapped to its corresponding *EQD2* value using the generated spline.

### 2.3 Statistical methods, parameter values, and implementation

The model and methods were implemented in Matlab (The Mathworks, Natick, USA).. The fitting methodology and statistical tools employed for this dataset are presented in detail in [5, 13]. The model was fitted to the clinical data using the maximum likelihood estimation (minimizing the function − ln *L*, where *L* is the likelihood), and confidence intervals for the best-fitting parameter values were computed using the profile likelihood methodology. The Akaike Information Criterion with sample size correction (*AIC*_c_) was used to evaluate model performance in comparison to other investigated models. Additionally, the *χ*^2^ test was used to evaluate the goodness of fit. To investigate the predictive performance of the model, a k-fold (*k*=2) cross-validation was employed. For this purpose, the datasets for LR and IR were split in two subsets; one of them was used for training and the other for validation. To ensure the representation of extreme hypofractionations, we included the constraint that each subset must contain at least two of the single-fraction treatments. This process was repeated 50 times to generate robust statistics.

Several parameters were fixed to values established in the literature. In the cell dynamics model, a value of *N*_tot_ = 10^10^ was assumed following [10], as this parameter was found to be non-significant for the primary objectives of the analysis. The tumor growth fractions (*GF*) and doubling times (*T*_D_) were taken from Zharinov *et al*. [14], who reported median growth fractions (Ki-67) of 0.083 and 0.0116 and doubling times of 34.83 and 30.34 months for tumors that can be classified as LR and IR, respectively.

Ljungkvist *et al*. [15] investigated the cell loss half-time (*T*_h_) for several types of cancer, reporting values in the range 17–49 h. Given the high degree of heterogeneity expected within the hypoxic compartment, a death rate corresponding to the lower limit of reported values was deemed appropriate; thus, *T*_h_ was set to 48 h as in [11]. Finally, the oxygen enhancement ratio for the hypoxic compartment (OER_*H*_) was set to 1.37 according to Chan *et al*. [16], and *T*_C_ was set to 48 h according to [17].

The value of *α* was fixed at 0.15 Gy^−1^ [13]. While *α* plays an important role in a mechanistic model like the MSK, particularly if fitting TCPs based on the viable number of cells and the LQ-Poisson formulation, the fits of a phenomenological logistic model like equation (15) are relatively insensitive to *α*. This is because a change in *D*_50_ can compensate for variations in *α* (this was confirmed by the sensitivity analysis).

Furthermore, the parameter space was constrained to ensure biologically and physically plausible solutions (with special emphasis on avoiding negative cell populations arising from Eqs. (13)-(14), and to improve computational convergence. The constraint OER*I ≥* OER*H* was enforced following [11]. The *α/β* ratio was constrained to relatively low values (*≤* 8 Gy as in [5], corresponding to 95% confidence intervals reported in [13]) to specifically check if reoxygenation can explain the response to extreme hypofractionation while being in agreement with the reported low *α/β* for prostate cancer.

Overall, the number of free parameters for model fitting was reduced to six: *α/β, k*_m_, OER*I* and *f*_pro_ (MSK model), along with *D*_50_ and *γ*_50_ (logistic TCP model). The constraints used during model fitting are shown in Table 2.

**Table 2:**
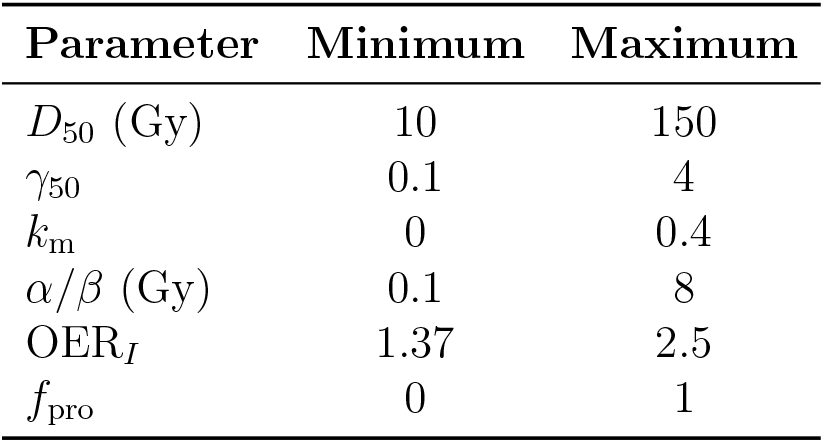
Optimization constraints and search space for the model parameters.

We used the Sobol methodology to investigate the parametric sensitivity of the model, computing first- and total-order Sobol sensitivity indices by using the methodology of Saltelli *et al*. [18]. Sensitivity was investigated in a hypercube covering ± 10% of the best-fitting values (or fixed values) sampling *N* = 10^5^ combinations of parameters. We used reparametrization to handle the constraints above imposed by Eqs. (13) and (14), defining *p*_1_ = *GF/f*pro and 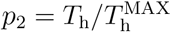, where 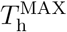 is the maximum value of *T*_h_ such that *QI ≥* 0 (therefore a function of *GF, f*_pro_, *T*_C_ and *T*_D_).

## 3 Results

In Figure 1, we present the best fits of the model to the dose-response dataset, independently for LR and IR patients. Single fraction schedules are highlighted in the figure. In Table 3, we present the best-fitting parameters for LR and IR, and the parameters describing the goodness-of-fit (− ln *L, AIC*_c_), and the *p* value obtained from the *χ*^2^ test. 95% CIs are also presented in the table. Because the computation of CI with the profile likelihood method is computationally demanding, we have restricted this computation, prioritizing the parameters of the MSK model (which characterize the mechanistic drivers of the response) over the phenomenological parameters of the logistic TCP.

**Table 3:** Best fits obtained with the MSK reoxygenation model to dose–response data for prostate cancer treated with HDR-BT, separated by risk (low, LR, and intermediate risk, IR). The table shows best fitting parameters, maximum likelihood and *AIC*_c_, and *p* value of the *χ*^2^ test. The symbols * and + are used to indicate that the parameter reached the edge of the constraint window, or that the parameter reaches the limit of positivity of cells given by equation (14).

|  | Low risk (LR) |  | Intermediate risk (IR) |  |
| --- | --- | --- | --- | --- |
|  | Values | 95% CI | Values | 95% CI |
| $\alpha/\beta$ [Gy] | 0.96 | [0.84, 8*] | 8.0* | [5.06, 8*] |
| $OER_I$ | 2.5* | [1.93, 2.5*] | 2.5* | [1.61, 2.5*] |
| $k_m$ | 0.0* | [0.0*, 0.40*] | 0.40* | [0.24, 0.4*] |
| $f_{pro}$ | 0.09 | [0.090<sup>+</sup>, 0.093] | 0.34 | [0.28, 0.48] |
| $D_{50}$ [Gy] | 25.11 | - | 26.73 | - |
| $\gamma_{50}$ | 0.66 | - | 1.16 | - |
| $-\ln L$ | 36.67 | - | 45.40 | - |
| $AIC_c$ | 91.34 | - | 108.06 | - |
| $p$ ($\chi^2$ test) | 1.0000 | - | 0.9998 | - |

**Figure 1:**
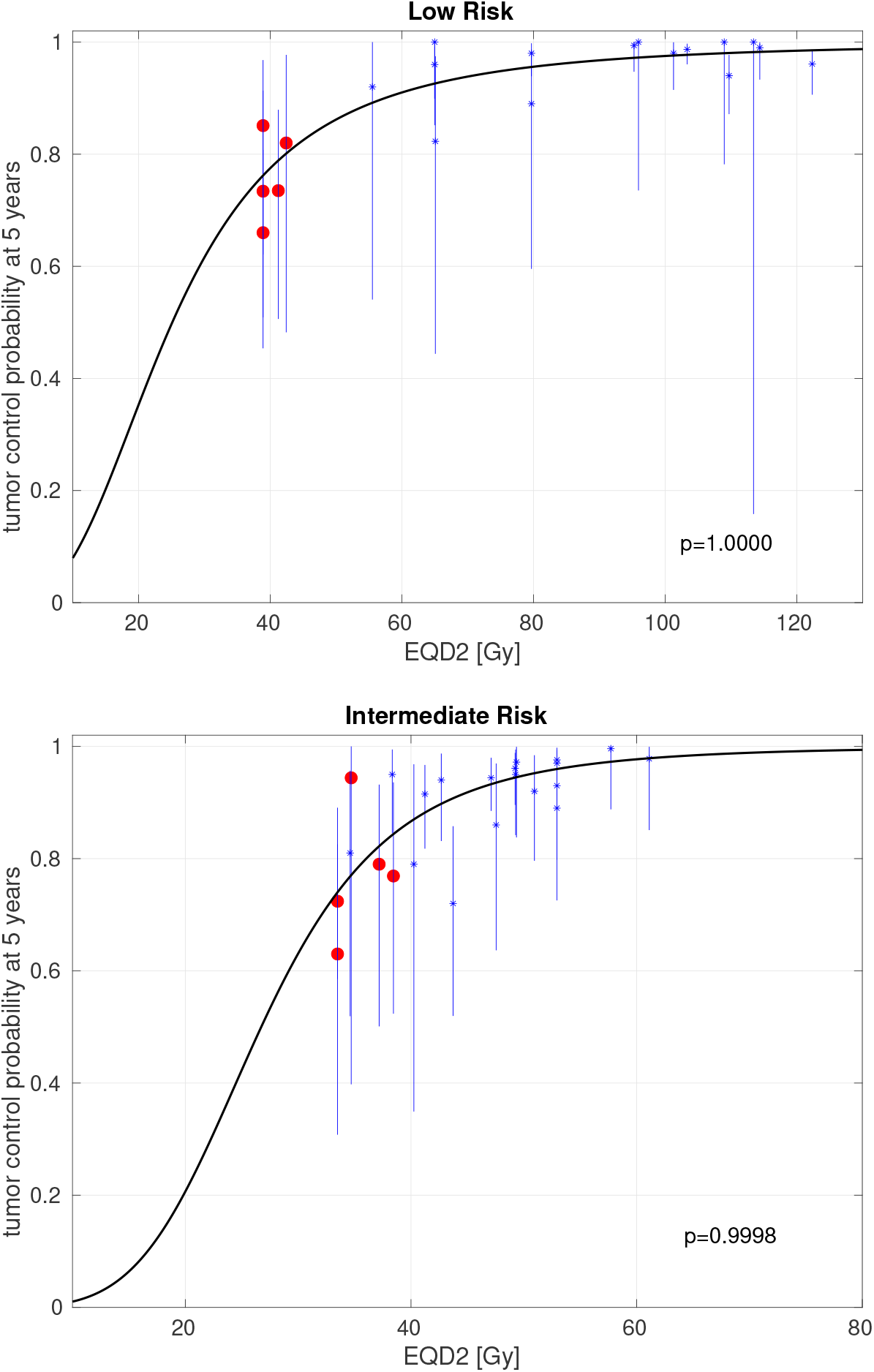
Best fits of the MSK reoxygenation model to dose–response data for prostate cancer treated with HDR-BT, separated in low risk (top panel) and intermediate risk (bottom panel). Clinical data (*) and 95% confidence intervals (bars), and modeled curves (solid lines). Single fraction schedules are highlighted as red circles. The *p* value of the *x*^2^ test is reported for each fit.

The results of the Sobol sensitivity analysis, first-order and total-order indices for each parameter, are presented in Table 4. The results of the *k*=2 cross-validation are presented in Table 5. In this case, we report the mean and standard deviation of the *p* values obtained from the *χ*^2^ test for the training and validation datasets (50 experiments).

**Table 4:** Parametric sensitivity analysis of the model: first-order (*S*) and total-order (*S*_*T*_) Sobol sensitivity indices.

| Parameters | LR |  | IR |  |
| --- | --- | --- | --- | --- |
| | $S$ | $S_T$ | $S$ | $S_T$ |
| $D_{50}$ | 0.042 | 0.088 | 0.293 | 0.909 |
| $\gamma_{50}$ | 0.006 | 0.020 | 0.000 | 0.387 |
| $p_1$ | 0.399 | 0.563 | 0.001 | 0.002 |
| $T_D$ | 0.000 | 0.000 | 0.000 | 0.000 |
| $T_C$ | 0.002 | 0.004 | 0.000 | 0.004 |
| $p_2$ | 0.266 | 0.422 | 0.000 | 0.000 |
| $k_m$ | 0.001 | 0.003 | 0.012 | 0.012 |
| $\alpha$ | 0.009 | 0.020 | 0.000 | 0.009 |
| $\alpha/\beta$ | 0.003 | 0.006 | 0.000 | 0.129 |
| $OER_I$ | 0.053 | 0.101 | 0.047 | 0.2213 |
| $OER_H$ | 0.000 | 0.000 | 0.000 | 0.000 |
| $f_{pro}$ | 0.002 | 0.004 | 0.000 | 0.003 |

**Table 5:** Results of the *k*=2 cross-validation: mean and standard deviation of the *p* values obtained from the *x*^2^ test for the training and validation datasets (50 experiments), for LR and IR.

| LR |  | IR |  |
| --- | --- | --- | --- |
| training | validation | training | validation |
| $0.917 \pm 0.064$ | $0.516 \pm 0.298$ | $0.873 \pm 0.114$ | $0.385 \pm 0.358$ |

## 4 Discussion

Several HDR-BT clinical trials have shown a significant loss of tumor control when treating prostate cancer with extremely hypofractionated protocols (∼ 20 Gy in a single fraction) [19, 20, 21, 22, 23, 24, 25], which is not consistent with the widely assumed low *α/β* ratio of prostate cancer and the LQ model. Among the causes that could be behind this behavior is the failure of the LQ model at large doses per pulse [7, 26], with some studies suggesting that it should be replaced by models like the LQL in this regime. Hypoxia/reoxygenation has also been suggested to play a role in the response of prostate cancer [8].

In a recent publication [5] we analyzed the response of prostate cancer to HDR-BT and found that the LQL model can fit the observed dose-response at large doses per pulse, while keeping the intrinsic *α/β* value low. This seemed to point out to a failure of the LQ model to fit data at large doses per fraction. In that work, we also performed a preliminary study of the role of reoxygenation with a simple reoxygenation model, but found that it could not fit the data as well as the LQL model. In this follow-up work we have performed a deeper investigation of the role of hypoxia and oxygenation in the response to HDR-BT ultra-fractionation by using a more complex and realistic model, which we call here the MSK model, originally introduced by Jeong *et al*. [10] and well validated to analyze clinical dose-response data [11].

We found that the LQ model and a low *α/β* value is consistent with the observed loss of tumor control of extremely hypofractionated protocols when reoxygenation is accounted for with the MSK model. Fits to LR and IR data showed *p*-values (*χ*^2^ test) of 1.0000 and 0.9998 respectively, which highlights the goodness of the fits. Moreover, the model was predictive as shown through a *k*=2 cross-validation: *p*-values of fits to the training dataset were 0.917 ± 0.064 and 0.873± 0.385 for LR and IR, respectively (average and standard deviation of 50 random experiments); *p*-values on the validation dataset were 0.516 ± 0.298 and 0.385± 0.358 for LR and IR, respectively.

These results were compared to those reported in [5] for the LQL model (notice that we used exactly the same dataset in both studies to facilitate the comparison). The *AIC*_c_ obtained in this work were 91.34 (LR) and 108.06 (IR), versus 89.25 (LR) and 103.41 (IR) obtained for the LQL. Therefore, the hypoxia/reoxygenation model was comparable to the LQL model for LR (Δ*AIC*c ≃ 2.1) and moderately inferior for IR (Δ*AIC*c ≃ 4.5), but differences in *AIC*_c_ above 10 are typically required to state the superiority of a given model [27]. Notice that the *AIC*_c_ penalizes the number of free parameters, and the MSK model inherently contains more parameters than the LQL model. However, because we have fixed several parameters according to the literature, the number of free parameters of both models is the same. Fits with the reoxygenation model were fairly superior to those with the LQ model when the *α/β* is constrained (*AIC*_c_ values of 97.30 and 117.95 for LR and IR, respectively).

Interestingly, the results of the best fits of Table 3 show a more important contribution of hypoxia for IR (34% of cells in the oxic proliferative compartment at baseline for IR versus 92% for LR), which agrees with experimental evidence [28], yet a faster reoxygenation for LR, shown by the value of *k*_m_. Despite the higher relative presence of cells in the intermediate/hypoxic compartments for IR, the optimizer was not able to find a better solution involving a faster reoxygenation (lower *k*_m_ value). Why this is the case it is not fully understood by the authors, but may be related to the model parameters that were constrained.

Studying the underlying mechanisms causing the significant loss of local control when delivering extremely hypofractionated HDR-BT seems of paramount importance to design effective clinical protocols. Our study suggests that reoxygenation can potentially explain these outcomes without departing from the standard LQ formalism, especially for LR data. Future clinical trials may shed new light on the response of prostate cancer to ultrahypofractionation. In particular, if hypoxia and reoxygenation are the primary drivers of this behavior rather than the mechanism behind the LQL response, an analysis of the role of pre-treatment tumor hypoxia (measured with functional imaging, for example) should show a far larger difference in tumor control between hypoxic and oxic tumors when treated with extreme hypofractionation than when treated with less radical fractionations. The role of hypoxia in the response to conventional fractionation is known, with Milosevic *et al*. [29] reporting a difference in local control of ∼7% between oxic and hypoxic tumors, but as far as we know a similar study has not been performed for extreme hypofractionation.

## 5 Conclusions

In this work, we have shown that the observed loss of tumor control in ultra-hypofractionated HDR-BT protocols for prostate cancer can be mechanistically explained by the LQ model with an intrinsic low *α/β* value by accounting for hypoxia and reoxygenation kinetics. If hypoxia and reoxygenation are indeed the drivers of this response, future clinical trials (prospective or retrospective) investigating the role of pre-treatment hypoxia status in tumor response may shed new light on this issue.

## Statements

### Data availability stattement

Code and supporting data are available from the Zenodo repository: https://doi.org/10.5281/zenodo.19923987.

